# Perceived association of mood and symptom severity in adults with mitochondrial diseases

**DOI:** 10.1101/2024.02.02.24302076

**Authors:** Catherine Kelly, Alex Junker, Kris Englestad, Michio Hirano, Caroline Trumpff, Martin Picard

## Abstract

Individuals with genetic mitochondrial diseases suffer from multisystemic symptoms that vary in severity from day-to-day and week-to-week, but the underlying causes of symptomatic fluctuations are not understood. Based upon observations that: i) patients and their families frequently report that stressful life events either trigger exacerbations of existing symptoms or the onset of new symptoms, ii) psychological states and stress hormones influence mitochondrial energy production capacity, and iii) epidemiological reports document a robust connection between traumatic/stressful life events and various neurologic disorders, we hypothesized that mitochondrial disease symptom severity may vary according to participant’s mood. To investigate this we administered the Stress, Health and Emotion Survey (SHES) in 70 adults (majority white (84%) cisgender women (83%), ages 18-74) with self-reported mitochondrial diseases (MELAS, 18%; CPEO, 17%; Complex I deficiency, 13%). Participants rated the severity of each of their symptom(s) over the past year on either good or bad days. On days marked by more stress, sadness and other negative emotions, some but not all symptoms were reported to be worse, including fatigue, exercise intolerance, brain fog, and fine motor coordination. By contrast, on days marked by happiness and calmness, participants reported these and other symptoms to be better, or less severe. Other symptoms including diminished sweating, hearing problems, and dystonia were in general unrelated to mood. Thus, some individuals living with mitochondrial diseases, at times perceive a connection between their mood and symptom severity. These preliminary associative results constitute an initial step towards developing more comprehensive models of the factors that influence the clinical course of mitochondrial diseases.

## Introduction

Mitochondrial diseases are a heterogenous group of disorders primarily caused by mutations or deletions in the mitochondrial DNA (mtDNA) or nuclear DNA (nDNA). Mitochondrial diseases are rare, child onset of mitochondrial disease ranges from 5-15 cases per 100,000, and adult onset is estimated to be 9.6 cases per 100,000 individuals (Emmanuele et al., 2022; Gorman et al., 2016). Current therapies are limited to symptom-specific treatment (e.g., blepharoplasty for ptosis) and preventative care (Tinker et al., 2021).The relative rarity of these conditions and the heterogeneity of symptoms contribute to a difficult diagnostic journey, with patients waiting on average 9 years for a diagnosis (Thompson et al., 2023). Individuals with the same mutation can have different organs affected, distinct clinical presentations, and a broad range of symptoms (Grier et al., 2018). The most common symptoms include fatigue, muscle weakness, gastrointestinal difficulties, ataxia, vision loss, and tremors (Gorman et al., 2016). While symptomatic heterogeneity and disease severity tend to differ according to specific mutations, and to be more severe in individuals with higher mtDNA mutation load (Grady et al., 2018), the course of mitochondrial disease is not linear, but varies over time (Dimond, 2013). Currently, we lack a full picture of the factors influencing the day-to-day and week-to-week symptomatic variations that patients experience.

Several common diseases are known to be triggered or exacerbated by psychosocial factors including mental stress (Hackett & Steptoe, 2017; Rosengren et al., 2004; Steptoe & Kivimaki, 2012; Thaker et al., 2006). Stress and other adverse psychological states produce physiological effects via multiple neuroendocrine pathways, where brain-derived signals cause the systemic release of glucocorticoids, catecholamines, and other neuroendocrine factors (O’Connor et al., 2021). Once in circulation, these potent hormones signal onto every organ system and target tissue (Basarrate et al., 2024), producing extensive immune and metabolic recalibrations (Cohen et al., 2012; Sterling, 2012). Thus, the metabolism of every organ system is hardwired (with receptors for circulating hormones or by nerve endings) to dynamically respond to one’s emotional state, or mood. This stress-metabolism cascade may explain how stressful experiences influence the symptomatology and/or pathogenesis of various somatic disorders, among which acute asthma (Chen & Miller, 2007) angina and coronary heart disease (Dimsdale, 2008; Fioranelli et al., 2018), and susceptibility to viral infections (Cohen et al., 1991) are best described. These data form the basis for a potentially modifiable stress-metabolism-disease cascade.

The existence of a stress-disease cascade is also supported by evidence in neurologic disorders. In comparison with the prevalence of adverse childhood experiences (ACEs) in the general population, ACEs were nearly twice as prevalent in outpatients with idiopathic neurologic and neuromuscular diseases (Mendizabal et al., 2022). High ACE scores were also associated with more frequent emergency visits and hospitalizations (Diaz et al., 2022; Okeson et al., 2022). Similarly, a meta-analysis including >150,000 adult participants showed that as the number of reported ACEs increased, the risk of primary headaches significantly increased, with both threat and deprivation traumas being associated with headache disorders (Sikorski et al., 2023), consistent with an effect of psychological factors on physical health. To our knowledge, whether psychosocial processes are associated with symptom severity in mitochondrial diseases has not been examined.

This study aimed to establish a connection between mood, and the severity of specific symptoms of mitochondrial diseases. We designed the *Stress, Health and Emotion Survey (SHES)* where participants reported the symptoms they experienced over the past year, and then rated the severity of each symptom on “good days” (described as happy, calm) and “bad” days (stressed, sad). Our findings exploring the perception of individuals with mitochondrial diseases extend existing literature on stress-disease experience to genetic mitochondrial disorders, calling for further research on potential strategies to improve patient care.

## Methods

### 1.1 Recruitment

We undertook an online survey of adults with self-reported mitochondrial disease and had previously agreed to be contacted for research studies. The survey was distributed through the North American Mitochondrial Disease Consortium (NAMDC) and United Mitochondrial Disease Foundation (UMDF) patient contact lists. This study was approved by the New York State Psychiatric Institute (NYSPI) [IRB protocol #8069].

### 1.2 Survey design and items

The SHES survey was designed to 1) record self-reported symptoms and their severity, and 2) assess whether their severity differs during “good” or “bad” days.

Our survey included 29 common mitochondrial disease symptoms compiled from the Newcastle Mitochondrial Disease Adult Scale (NMDAS)(Schaefer et al., 2006), Composite Autonomic Symptom Score-31 (COMPASS-31) (Sletten et al., 2012; Suarez et al., 1999), Columbia Neurological Scale (CNS) (Kaufmann et al., 2009; Kaufmann et al., 2011; Kaufmann et al., 2004), and input from a mitochondrial medicine specialist (author M.H.). Clinical symptom names were translated into non-scientific terms (e.g., “myoclonus” into “body/limb jerks”; full list of terms that differ between the survey and displayed figures is in **Appendix 1**).

Our survey first asked participants to check all symptoms they experienced over the past year. The survey then prompted participants to think about “good” emotional days (defined with positive words from the circumplex model of affect, “happy” and “calm”) and estimate how many good days they had over the past year. Participants were then asked to think about the severity of each endorsed symptom differed on good as compared to normal days, and rate this using a 7-point Likert scale ranging from “much worse” to “much better,” with 4 meaning “no difference.” After participants had rated symptom severity on good days, they were asked to repeat this process for “bad” emotional days (described with negative words from the circumplex model, “stressed” and “sad”). Patients reported on average 54% good days (range 10-98%, median 50%) and 42% bad days (range 5-90%, median 40%) in their previous year.

Sex was measured using self-selection of female, male, intersex, or prefer not to disclose. Gender was assessed with a check-all-that-apply question including woman, man, agender, genderqueer, gender-fluid, non-binary, questioning, transgender, trans man, trans woman, other, and prefer not to disclose. Race/ethnicity was measured with a check-all-that-apply question including American Indian, Asian, Black, Hispanic, Hawaiian and Pacific Islander, white, and/or prefer not to disclose. We did not collect any information on current medications, other diagnoses or mutations, insurance coverage status, class, exposure to stress, resources/support for mitochondrial disease, or challenges in participants’ daily lives. Notably, our survey cannot speak to the directionality of the relationship between symptom severity and mood - i.e., whether lessened symptoms led to days being more pleasant, or whether emotionally challenging days worsened symptoms.

At the end of the survey, participants were offered an open text box to contribute comments related to the survey. To gauge respondent perceptions of our central question and methods, we analyzed participant comments for their feedback on our methods and central question. Responses were coded and emergent themes are discussed below as guidance for future studies.

### 1.3 Statistical analyses

Participants were excluded if they did not complete the entire questionnaire (n=2) or were confused about the wording (as they noted in a comment, n=1). If participants completed the survey multiple times, the first survey was used, and the repeats were removed. Symptom-specific analyses were only run when n>3 people reported the symptom. “Good day” and “bad day” scores per symptom were calculated as the average severity score across all participants. The associations between average symptom severity and age or number of reported symptoms were assessed using Spearman’s rho correlation. The difference of average symptom severity between good and bad days was analyzed using a one-sample T-test. All statistical analyses were performed using Prism (version 9.5.1) or Excel (version 16.74).

## Results

### 2.1 Patient characteristics

A total of 70 participants with a self-reported mitochondrial disease completed the online survey. Participants were majority white (84%) cisgender women (83%), aged 18-74 years (median 50.5) (**Table 1**). 62 participants (87%) could list their specific clinical diagnosis. Mitochondrial Encephalomyopathy, Lactic Acidosis, and Stroke-like episodes (MELAS) was most frequently reported (18%), followed by Chronic Progressive External Ophthalmoplegia (CPEO) (17%), and Complex I deficiency (13%; **Figure 1A**, full list in **Table 1**). Most participants (n=60, 85%) did not know their genetic diagnosis.

**Table 1.** Participant characteristics.

| Variable <sup>a</sup> | Sample |
| --- | --- |
| N | 71 |
| Age, years (mean; range) | 48.1 (14.3) 19-74 |
| Sex, female | 59 (83.1) |
| Race/ethnicity |  |
| White | 60 (84.5) |
| Black | 2 (2.8) |
| Asian | 2 (2.8) |
| Hispanic or Latino | 2 (2.8) |
| Multiple | 2 (2.8) |
| Unknown | 2 (2.8) |
| American Indian or<br>Alaskan Native | 1 (1.4) |
| Symptom Number | 13.17 (5.57) |
| Reported clinical diagnosis |  |
| MELAS | 13 (18.3) |
| CPEO | 12 (16.9) |
| Complex I deficiency | 9 (12.8) |
| Complex III deficiency | 8 (11.3) |
| Complex IV deficiency / COX Deficiency | 6 (8.5) |
| Mitochondrial cytopathy | 6 (8.5) |
| Mitochondrial DNA depletion | 6 (8.5) |
| POLG mutations | 6 (8.5) |
| Co-Enzyme Q10 deficiency | 5 (7.0) |
| KSS | 5 (7.0) |
| Complex V deficiency | 4 (5.6) |
| Mitochondrial encephalopathy | 4 (5.6) |
| LHON | 3 (4.2) |
| Creatine deficiency syndromes | 3 (4.2) |
| Complex II deficiency | 3 (4.2) |
| Lactic acidosis | 2 (2.8) |
| Beta-oxidation defects | 1 (1.4) |
| Carnitine-Acyl-Carnitine deficiency | 1 (1.4) |
| MERRF | 1 (1.4) |
| MNGIE | 1 (1.4) |
| SCAD | 1 (1.4) |
| Unknown (Mitochondrial Cytopathy) | 9 (12.7) |
| Reported genetic diagnosis |  |
| m.3243A>G | 5 (7.0) |
| Single deletion | 4 (5.6) |
| m.14484A>G | 1 (1.4) |
| m.15257G>A | 1 (1.4) |
| Unknown | 60 (84.5) |

<sup>a</sup> Data presented as means (standard deviation) or n (%)

**Figure 1.**
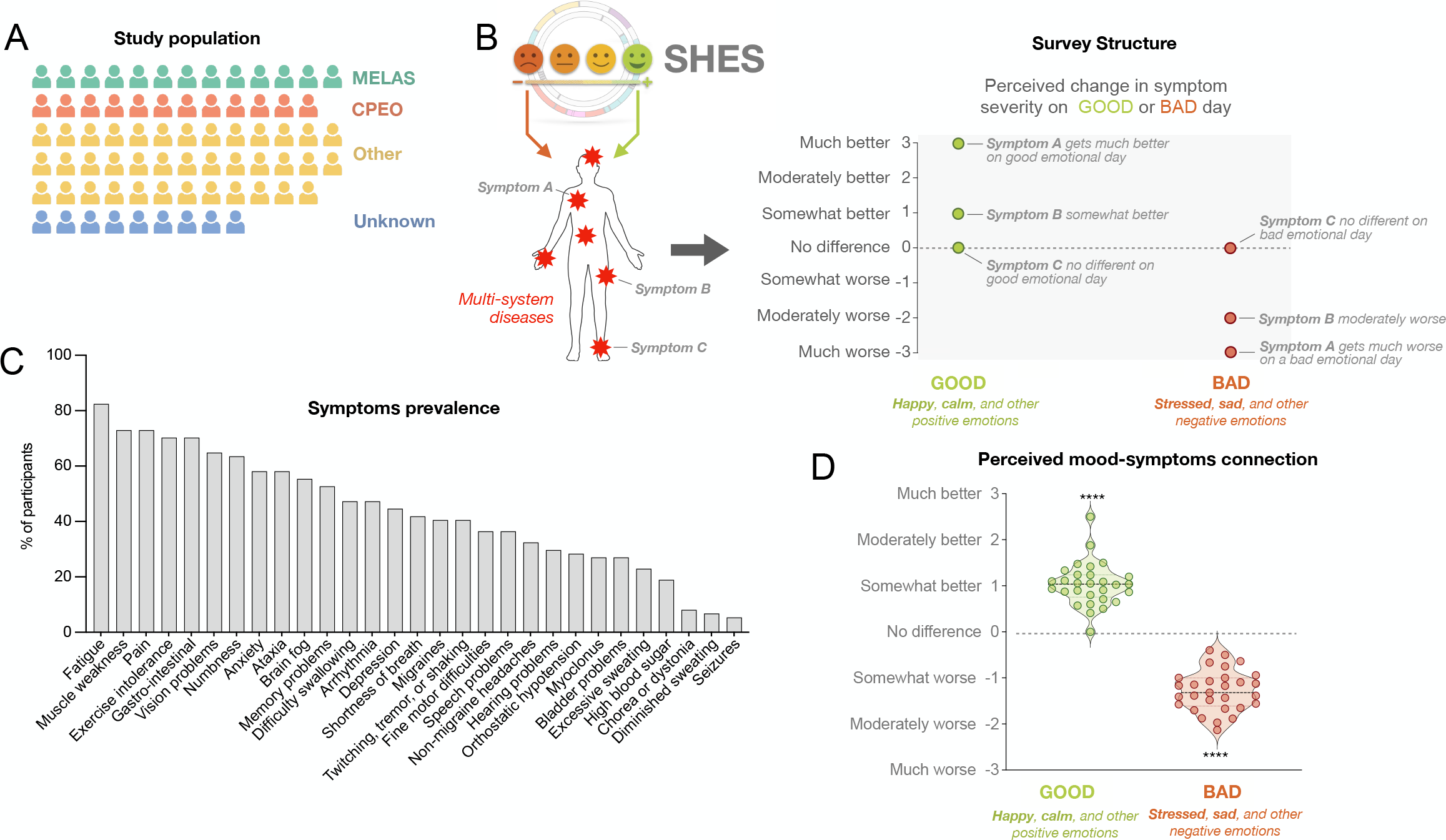
Survey administered to quantify perceived severity of mitochondrial disease symptoms according do mood. (**A**) Depiction of the heterogenous clinical diagnoses among study participants. (**B**) The Stress, Health and Emotions Survey (SHES) first asked participants to select the symptoms they have experienced in the past year. Next, participants indicated on a 7-point Likert scale ranging from from “much worse” to “much better” the extent to which each symptom differs on days marked by feelings of happiness and calmness (*good days*), and how the same symptom differs on days marked by feelings of stress and sadness (*bad days*). (**C**) Percentage of sample population (n=70) reporting each symptom. Fatigue (82.4%), muscle weakness (73.0%), and pain (73.0%) were most prevalent. (**D**) Difference in perceived symptoms severity computed as a score reflecting the average across all participants for each symptom. Datapoints far away from the dotted line (null hypothesis: no difference) are symptoms perceived to be more strongly to related to mood, in either direction. Bar in violin plots represent the mean of all symptoms. **** p<0.0001, one-sample t test against null hypothesis (y=0, no difference).

### 2.2 Symptom prevalence

Participants endorsed on average 13 symptoms (range: 2-24, median 13). Aligning with previous literature (Ameele, 2020; Gorman et al., 2015; Kanungo et al., 2018) the most commonly endorsed symptoms were fatigue (82% of participants), muscle weakness (73%), and pain (73%). The most rarely endorsed symptoms were seizures (6%), diminished sweating (7%), and involuntary muscle jerks (8%) (**Figure 1C**).

### 2.3 Perceived differences in symptom severity by positive and negative mood

On average, patients reported that most symptoms were better (i.e., less severe) on days marked by more happy and calm feelings, or “good” emotional days (p<0.0001). On the other hand, most symptoms were generally worse on days with more feelings of stress, sadness and other negative emotions, or “bad” emotional days (p<0.0001, **Figure 1D**). Across all symptoms, there was a significant difference in perceived symptom severity between good and bad days (p<0.0001). These effects did not significantly vary by number of symptoms reported (Spearman r=0.002, p=0.99) nor by age (r=-0.10, p=0.30).

Symptoms that were significantly better on *good days* included seizures, depression, anxiety, fatigue, fine motor coordination, and non-migraine headaches (all p<0.05). Symptoms that were significantly worse on *bad days* included fatigue, depression, anxiety, exercise intolerance, brain fog, and fine motor coordination (all p<0.05) (**Figure 2**). The severity reported by each participant, by symptom, are shown in **Supplemental Figure S1**.

**Figure 2.**
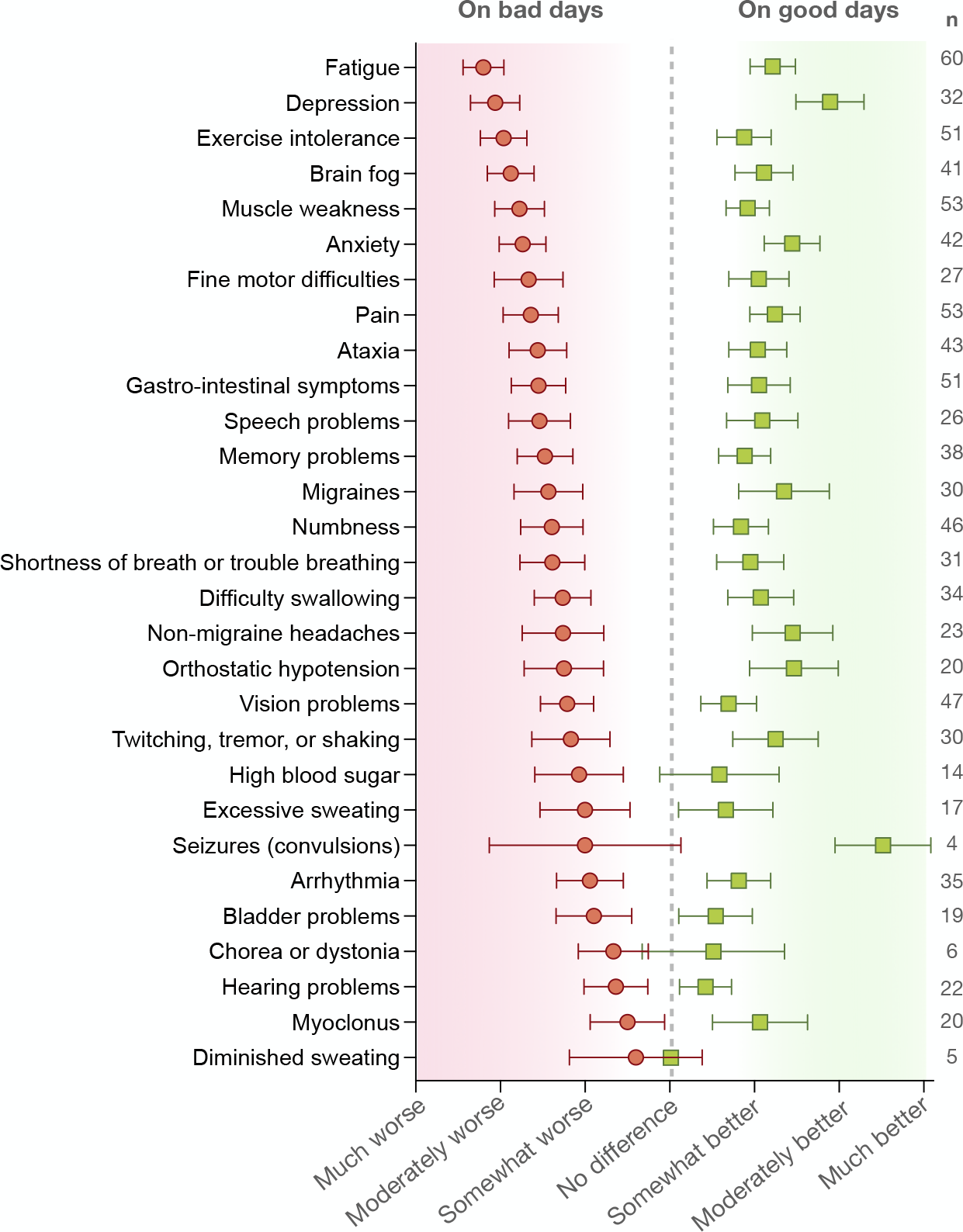
Participant-rated difference in perceived symptom severity based on good or bad emotional days. Average difference in symptom severity ranked from most (top) to least (bottom) associated with mood (stress, sad and other negative feelings) on bad days, and the corresponding score for good days (happy, calm and other positive feelings). The number of participants endorsing each symptom (n) is shown on the right. Datapoints are average change score with 95% confidence interval across all participants. See Supplemental Figure S1 for details by symptom, with datapoints for each participant.

The specificity of symptoms reported to be related to daily mood provides some evidence that participants actively engaged with this survey and did not universally conflate physical and psychological states. The severity of some symptoms showed no association with mood, including diminished sweating, hearing problems, chorea/dystonia, and myoclonus. The lack of perceived association between mood and certain symptoms increases the probability that the consistent effects reported for the symptoms above are specific and valid.

Finally, perceived differences in symptom severity on “good” and “bad” days were similar between clinical groups (CPEO, MELAS, and other clinical diagnoses) (**Supplemental Figure S2**). All clinical groups rated exercise intolerance and fatigue among the symptoms that differ the most on bad days and numbness and vision problems among the symptoms that differ the least on bad days.

## Discussion

The aim of this study was to explore how adults with mitochondrial diseases perceive the link between their daily mood and their symptoms. The signs and symptoms of mitochondrial diseases do not progress linearly. They fluctuate over time, at times worsening, and at times improving for unknown reasons. Since the underlying gene defect does not dynamically amplify or regress over days and weeks, whereas symptoms do vary across these time-scales, other disease-modifying factors are likely at play. Building from converging literatures demonstrating that multiple organ systems implicated in mitochondrial disease symptomatology are regulated by stress hormones released in response to psychological stress (Bodine & Furlow, 2015; Tsigos et al., 2000), and that mitochondrial respiratory capacity is sensitive to stress hormones and mood (Picard & McEwen, 2018; Picard et al., 2018; Psarra & Sekeris, 2011), mood emerges a potential modifier of mitochondrial disease pathophysiology.

The results suggest some rather specific mood-symptom associations. Some symptoms were consistently ranked as unchanging or stable across good and bad emotional days. This includes hearing problems, diminished sweating, jerky muscle movements, arrhythmia, and vision problems. The majority of these symptoms are possibly more directly caused by fixed structural lesions, such as vision that can be impaired by the optic neuropathy or pigmentary retinopathy (Zeviani & Carelli, 2021). In contrast, some symptoms were ranked as more variable including fatigue, depression, brain fog, fine motor coordination, ataxia, and gastrointestinal symptoms. These are either inherently subjective or physiological in nature. Some symptoms, like exercise intolerance, were ranked as worse on bad days, but on aggregate not significantly different on good ones, suggesting potentially specific psychobiological effects of positive and negative mood. Unsurprisingly, subjective symptoms such as pain and anxiety are both reported to be better on good days, and worse on days when individuals with mitochondrial diseases feel stressed or sad. If interpreted in the opposite and (at least) equally likely directionality, patients indicated that good days are ones with less pain, while bad days are ones where they are exhausted and cannot tolerate movement.

Patients with mitochondrial disease have previously indicated that they would like doctors’ attention on the emotional sides of living with their disease (Zilber, 2020), including asking how they are doing during appointments (Valverde et al., 2022). In one qualitative study, a participant voiced that, in her two decades of being diagnosed with mitochondrial disease, the research interview was the first time she had been asked questions about her emotions (Valverde et al., 2022). This is particularly worrisome as the aspects of mitochondrial disease that physicians expect to be most burdensome often do not line up with the experience of patients or their family members (Koene et al., 2013). When some mitochondrial disease patients were asked what metrics they most want to improve, mental health was the second most commonly endorsed answer, only after energy level (Karaa et al., 2019). The diagnostic and prognostic uncertainty of mitochondrial diseases alone can be distressing, as can living with symptoms that unpredictably limit what one can do in a day (Valverde et al., 2022). Additionally, 33% of patients said that their mental health was negatively impacted by the limitations they faced with their healthcare provider (Karaa et al., 2019). There is an unmet need to address the psychological aspects of living with mitochondrial diseases. In addition to improving well-being and patient satisfaction, if the results of this survey are validated in future research, addressing the psychological needs of patients could contribute to the clinical management of symptoms.

Several participants commented on the stress-disease connection in relation to mitochondrial diseases. For example, some participants believed that the connection between mood and symptom severity may be more relevant for some people than others, or only be relevant for a period of a person’s life (e.g., during a major stressor). One participant said their disease was triggered by a well-defined period of intense stress, whereas others stated their mitochondrial disease symptoms and moods are entirely independent of each other. This heterogeneity in perspectives aligns well with the range of responses to the main mood-symptom SHES question, as well as with previous findings that research questions about stress/mental health in mitochondrial disease research are polarizing, with some patients viewing it as essential and some as irrelevant (Thomas et al., 2022). Moreover, some participants noted that the SHES survey could not fully capture how they experience their symptoms and mood interacting. One participant described their symptom severity as a moving target as their disease changes over time. Participants also pointed out our failure to ask about coping strategies or financial barriers to care; future research should try to capture the material reality of participants and the active strategies they are using to live with their conditions. Future research would also gain by using mixed-methods designs that can better capture the richness of individual experiences.

This study has several limitations. Retrospective designs rely on recall and are therefore associated with recall bias (Ottenstein, 2020; Ready R.E., 2005; Sato & Kawahara, 2011; Van den Bergh & Walentynowicz, 2016). Importantly, our survey cannot speak to the directionality of the association between mood and symptom severity. The SHES survey explicitly asked participants to rank how their symptoms differed on good or bad days, presupposing that days are good or bad independent of symptoms and the limitations they impose on daily activity, which some participants noted as inaccurate. The sample size in this cohort is also small, but the range of symptoms, age, and clinical diagnoses match the one of larger cohorts (Barca et al., 2020)

Establishing psychobiological modifiers of mitochondrial diseases requires several sequential pieces of evidence. A survey such as SHES is the first and weakest piece of evidence. These preliminary results now call for studies using specific operationalizations of psychological and biological stress markers. Experimental laboratory stress paradigms can also be used to elicit and provide more precise quantification of the psychobiological reactivity and recovery to challenges (Kirschbaum et al., 1993). Measures of cumulative stress, such as allostatic load (Bobba-Albes, 2022) or possibly epigenetic clocks (Kabacik et al., 2022), also could be used to quantify the biological toll of chronic stress across organ systems. Combined, these approaches would help to delineate how psychological experiences may ‘get under the skin’ and contribute to the course of neuromuscular disorders through biologically plausible endocrine, metabolic, immune, or other pathways. In parallel, the reciprocal connection between mood and symptoms would be better mapped by qualitative methods. Thus, while preliminary, these results call for prospective, mixed-methods research using repeated sampling to quantifiably define the interaction between psychosocial factors and neurometabolic processes linked to mitochondrial biology and symptomatology. This research could reveal modifiable drivers of disease risk, and perhaps identify factors that confer resilience among certain patients with mitochondrial diseases.

## Data Availability

All data produced in the present study are available upon reasonable request to the authors

## Acknowledgments

This work was supported by NIH grant R01MH122706, U54NS078059, the Wharton Fund and the Baszucki Brain Fund. The authors would like to thank the patients for their participation.

## Data sharing statement

Anonymized raw data may be made available upon request to the corresponding author, MP.

## Supplemental information

- Supplemental Figure S1
- Supplemental Figure S2
- Appendix 1

## Appendix 1. Changes in symptom wording between the survey and the presented figures

**Appendix 1.** Changes in symptom wording between the survey and the presented figures.
| Questionnaire wording | Figure wording |
| --- | --- |
| Abnormal movements (chorea or dystonia) | Chorea or dystonia |
| Bladder problems (such as difficulty passing urine, loss of bladder control) | Bladder problems |
| Arrhythmia or other heart symptoms (such as chest pain, palpitations, a fluttering in your chest) | Arrhythmia |
| Numbness, tingling, burning pain in my hands, feet, or both | Numbness |
| Muscle weakness (such as difficulty standing up, climbing stairs) | Muscle weakness |
| Trouble with balance (incoordination or ataxia) | Ataxia |
| Gastro-intestinal symptoms (such as nausea, vomiting, constipation, diarrhea) | Gastro-intestinal symptoms |
| Pain (such as joint pain, back pain, muscle pain) | Pain |
| Body limbs/jerks (myoclonus) | Myoclonus |
| Finger/hand motor coordination difficulties | Fine motor difficulties |
| Seizures (convulsions) | Seizures |
| Fainting or feeling faint when standing | Orthostatic hypotension |

## References

Ameele, J. V. D., Fuge, J., Pitceathly, R.D.S., Berry, S., McIntyre Z., Hanna, M.G., Lee, M., Chinnery, P.F. . (2020). Chronic pain is common in mitochondrial disease. Neuromuscular Disorders, 30(5), 413–419. 10.1016/j.nmd.2020.02.017

Barca, E., Long, Y., Cooley, V., Schoenaker, R., Emmanuele, V., DiMauro, S., Cohen, B. H., Karaa, A., Vladutiu, G. D., Haas, R., Van Hove, J. L. K., Scaglia, F., Parikh, S., Bedoyan, J. K., DeBrosse, S. D., Gavrilova, R. H., Saneto, R. P., Enns, G. M., Stacpoole, P. W., … Hirano, M. (2020). Mitochondrial diseases in North America: An analysis of the NAMDC Registry. Neurol Genet, 6(2), e402. 10.1212/NXG.0000000000000402

Basarrate, S., Monzel, A. S., Smith, J., Marsland, A., Trumpff, C., & Picard, M. (2024). Glucocorticoid and adrenergic receptor distribution across human organs and tissues: a map for stress transduction. Psychosom Med. 10.1097/PSY.0000000000001275

Bobba-Albes, N. J. R.P,; Picard, M. (2022). The energetic cost of allostasis and allostatic load. Psychoneuroendocrinology, 146. 10.1016/j.psyneuen.2022.105951

Bodine, S. C., & Furlow, J. D. (2015). Glucocorticoids and Skeletal Muscle. Adv Exp Med Biol, 872, 145–176. 10.1007/978-1-4939-2895-8_7

Chen, E., & Miller, G. E. (2007). Stress and inflammation in exacerbations of asthma. Brain Behav Immun, 21(8), 993–999. 10.1016/j.bbi.2007.03.009

Cohen, S., Janicki-Deverts, D., Doyle, W. J., Miller, G. E., Frank, E., Rabin, B. S., & Turner, R. B. (2012). Chronic stress, glucocorticoid receptor resistance, inflammation, and disease risk. Proc Natl Acad Sci U S A, 109(16), 5995–5999. 10.1073/pnas.1118355109

Cohen, S., Tyrrell, D. A., & Smith, A. P. (1991). Psychological stress and susceptibility to the common cold. N Engl J Med, 325(9), 606–612. 10.1056/NEJM199108293250903

Diaz, R., Walker, R. J., Lu, K., Weston, B. W., Young, N., Fumo, N., & Hilgeman, B. (2022). The relationship between adverse childhood experiences, the frequency and acuity of emergency department utilization and primary care engagement. Child Abuse Negl, 124, 105479. 10.1016/j.chiabu.2021.105479

Dimond, R. (2013). Patient and family trajectories of mitochondrial disease: diversity, uncertainty and genetic risk. Life Sciences, Society and Policy, 9. 10.1186/2195-7819-9-2

Dimsdale, J. E. (2008). Psychological stress and cardiovascular disease. J Am Coll Cardiol, 51(13), 1237–1246. 10.1016/j.jacc.2007.12.024

Emmanuele, V., Ganesh, J., Vladutiu, G., Haas, R., Kerr, D., Saneto, R. P., Cohen, B. H., Van Hove, J. L. K., Scaglia, F., Hoppel, C., Rosales, X. Q., Barca, E., Buchsbaum, R., Thompson, J. L., DiMauro, S., Hirano, M., & North American Mitochondrial Disease, C. (2022). Time to harmonize mitochondrial syndrome nomenclature and classification: A consensus from the North American Mitochondrial Disease Consortium (NAMDC). Mol Genet Metab, 136(2), 125–131. 10.1016/j.ymgme.2022.05.001

Fioranelli, M., Bottaccioli, A. G., Bottaccioli, F., Bianchi, M., Rovesti, M., & Roccia, M. G. (2018). Stress and Inflammation in Coronary Artery Disease: A Review Psychoneuroendocrineimmunology-Based. Front Immunol, 9, 2031. 10.3389/fimmu.2018.02031

Gorman, G. S., Chinnery, P. F., DiMauro, S., Hirano, M., Koga, Y., McFarland, R., Suomalainen, A., Thorburn, D. R., Zeviani, M., & Turnbull, D. M. (2016). Mitochondrial diseases. Nat Rev Dis Primers, 2, 16080. 10.1038/nrdp.2016.80

Gorman, G. S., Elson, J. L., Newman, J., Payne, B., McFarland, R., Newton, J. L., & Turnbull, D. M. (2015). Perceived fatigue is highly prevalent and debilitating in patients with mitochondrial disease. Neuromuscul Disord, 25(7), 563–566. 10.1016/j.nmd.2015.03.001

Grady, J. P., Pickett, S. J., Ng, Y. S., Alston, C. L., Blakely, E. L., Hardy, S. A., Feeney, C. L., Bright, A. A., Schaefer, A. M., Gorman, G. S., McNally, R. J., Taylor, R. W., Turnbull, D. M., & McFarland, R. (2018). mtDNA heteroplasmy level and copy number indicate disease burden in m.3243A>G mitochondrial disease. EMBO Mol Med, 10(6). 10.15252/emmm.201708262

Grier, J., Hirano, M., Karaa, A., Shepard, E., & Thompson, J. L. P. (2018). Diagnostic odyssey of patients with mitochondrial disease: Results of a survey. Neurol Genet, 4(2), e230. 10.1212/NXG.0000000000000230

Hackett, R. A., & Steptoe, A. (2017). Type 2 diabetes mellitus and psychological stress - a modifiable risk factor. Nat Rev Endocrinol, 13(9), 547–560. 10.1038/nrendo.2017.64

Kabacik, S., Lowe, D., Fransen, L., Leonard, M., Ang, S. L., Whiteman, C., Corsi, S., Cohen, H., Felton, S., Bali, R., Horvath, S., & Raj, K. (2022). The relationship between epigenetic age and the hallmarks of aging in human cells. Nat Aging, 2(6), 484–493. 10.1038/s43587-022-00220-0

Kanungo, S., Morton, J., Neelakantan, M., Ching, K., Saeedian, J., & Goldstein, A. (2018). Mitochondrial disorders. Ann Transl Med, 6(24), 475. 10.21037/atm.2018.12.13

Karaa, A., Goldstein, A., Balcells, C., Mann, K., Stanley, L., Yeske, P. E., & Parikh, S. (2019). Primary mitochondrial disease in the US: Data from patients and physicians’ perspective on health care delivery. Data Brief, 25, 104343. 10.1016/j.dib.2019.104343

Kaufmann, P., Engelstad, K., Wei, Y., Kulikova, R., Oskoui, M., Battista, V., Koenigsberger, D. Y., Pascual, J. M., Sano, M., Hirano, M., DiMauro, S., Shungu, D. C., Mao, X., & De Vivo, D. C. (2009). Protean phenotypic features of the A3243G mitochondrial DNA mutation. Arch Neurol, 66(1), 85–91. 10.1001/archneurol.2008.526

Kaufmann, P., Engelstad, K., Wei, Y., Kulikova, R., Oskoui, M., Sproule, D. M., Battista, V., Koenigsberger, D. Y., Pascual, J. M., Shanske, S., Sano, M., Mao, X., Hirano, M., Shungu, D. C., Dimauro, S., & De Vivo, D. C. (2011). Natural history of MELAS associated with mitochondrial DNA m.3243A>G genotype. Neurology, 77(22), 1965–1971. 10.1212/WNL.0b013e31823a0c7f

Kaufmann, P., Shungu, D. C., Sano, M. C., Jhung, S., Engelstad, K., Mitsis, E., Mao, X., Shanske, S., Hirano, M., DiMauro, S., & De Vivo, D. C. (2004). Cerebral lactic acidosis correlates with neurological impairment in MELAS. Neurology, 62(8), 1297–1302. 10.1212/01.wnl.0000120557.83907.a8

Kirschbaum, C., Pirke, K. M., & Hellhammer, D. H. (1993). The ‘Trier Social Stress Test’--a tool for investigating psychobiological stress responses in a laboratory setting. Neuropsychobiology, 28(1-2), 76–81. 10.1159/000119004

Koene, S., Jansen, M., Verhaak, C. M., De Vrueh, R. L., De Groot, I. J., & Smeitink, J. A. (2013). Towards the harmonization of outcome measures in children with mitochondrial disorders. Dev Med Child Neurol, 55(8), 698–706. 10.1111/dmcn.12119

Mendizabal, A., Nathan, C. L., Khankhanian, P., Anto, M., Clyburn, C., Acaba-Berrocal, A., Breen, L., & Dahodwala, N. (2022). Adverse Childhood Experiences in Patients With Neurologic Disease. Neurol Clin Pract, 12(1), 60–67. 10.1212/CPJ.0000000000001134

O’Connor, D. B., Thayer, J. F., & Vedhara, K. (2021). Stress and Health: A Review of Psychobiological Processes. Annu Rev Psychol, 72, 663–688. 10.1146/annurev-psych-062520-122331

Okeson, K., Reid, C., Mashayekh, S., Sonu, S., Moran, T. P., & Agarwal, M. (2022). Adverse Childhood Experiences and Healthcare Utilization of Children in Pediatric Emergency Departments. J Pediatr, 240, 206–212. 10.1016/j.jpeds.2021.09.016

Ottenstein, C., Lischetzke, T. (2020). Recall bias in emotional intensity ratings: investigating person-level and event-level predictors. Motivation and Emotion, 44, 464–473. 10.1007/s11031-019-09796-4

Picard, M., & McEwen, B. S. (2018). Psychological Stress and Mitochondria: A Systematic Review. Psychosom Med, 80(2), 141–153. 10.1097/PSY.0000000000000545

Picard, M., Prather, A. A., Puterman, E., Cuillerier, A., Coccia, M., Aschbacher, K., Burelle, Y., & Epel, E. S. (2018). A Mitochondrial Health Index Sensitive to Mood and Caregiving Stress. Biol Psychiatry, 84(1), 9–17. 10.1016/j.biopsych.2018.01.012

Psarra, A. M., & Sekeris, C. E. (2011). Glucocorticoids induce mitochondrial gene transcription in HepG2 cells: role of the mitochondrial glucocorticoid receptor. Biochim Biophys Acta, 1813(10), 1814–1821. 10.1016/j.bbamcr.2011.05.014

Ready R.E. W. M.I., Jones K.M. (2005). How happy have you felt lately? Two diary studies of emotion recall in older and younger adults. Cognition and Emotion, 21(4), 728–757. 10.1080/02699930600948269

Rosengren, A., Hawken, S., Ounpuu, S., Sliwa, K., Zubaid, M., Almahmeed, W. A., Blackett, K. N., Sitthiamorn, C., Sato, H., Yusuf, S., & investigators, I. (2004). Association of psychosocial risk factors with risk of acute myocardial infarction in 11119 cases and 13648 controls from 52 countries (the INTERHEART study): case-control study. Lancet, 364(9438), 953–962. 10.1016/S0140-6736(04)17019-0

Sato, H., & Kawahara, J. (2011). Selective bias in retrospective self-reports of negative mood states. Anxiety Stress Coping, 24(4), 359–367. 10.1080/10615806.2010.543132

Schaefer, A. M., Phoenix, C., Elson, J. L., McFarland, R., Chinnery, P. F., & Turnbull, D. M. (2006). Mitochondrial disease in adults: a scale to monitor progression and treatment. Neurology, 66(12), 1932–1934. 10.1212/01.wnl.0000219759.72195.41

Sikorski, C., Mavromanoli, A. C., Manji, K., Behzad, D., & Kreatsoulas, C. (2023). Adverse Childhood Experiences and Primary Headache Disorders: A Systematic Review, Meta-analysis, and Application of a Biological Theory. Neurology, 101(21), e2151–e2161. 10.1212/WNL.0000000000207910

Sletten, D. M., Suarez, G. A., Low, P. A., Mandrekar, J., & Singer, W. (2012). COMPASS 31: a refined and abbreviated Composite Autonomic Symptom Score. Mayo Clin Proc, 87(12), 1196–1201. 10.1016/j.mayocp.2012.10.013

Steptoe, A., & Kivimaki, M. (2012). Stress and cardiovascular disease. Nat Rev Cardiol, 9(6), 360–370. 10.1038/nrcardio.2012.45

Sterling, P. (2012). Allostasis: a model of predictive regulation. Physiol Behav, 106(1), 5–15. 10.1016/j.physbeh.2011.06.004

Suarez, G. A., Opfer-Gehrking, T. L., Offord, K. P., Atkinson, E. J., O’Brien, P. C., & Low, P. A. (1999). The Autonomic Symptom Profile: a new instrument to assess autonomic symptoms. Neurology, 52(3), 523–528. 10.1212/wnl.52.3.523

Thaker, P. H., Han, L. Y., Kamat, A. A., Arevalo, J. M., Takahashi, R., Lu, C., Jennings, N. B., Armaiz-Pena, G., Bankson, J. A., Ravoori, M., Merritt, W. M., Lin, Y. G., Mangala, L. S., Kim, T. J., Coleman, R. L., Landen, C. N., Li, Y., Felix, E., Sanguino, A. M., … Sood, A. K. (2006). Chronic stress promotes tumor growth and angiogenesis in a mouse model of ovarian carcinoma. Nat Med, 12(8), 939–944. nm1447 [pii] 10.1038/nm1447

Thomas, R. H., Hunter, A., Butterworth, L., Feeney, C., Graves, T. D., Holmes, S., Hossain, P., Lowndes, J., Sharpe, J., Upadhyaya, S., Varhaug, K. N., Votruba, M., Wheeler, R., Staley, K., & Rahman, S. (2022). Research priorities for mitochondrial disorders: Current landscape and patient and professional views. J Inherit Metab Dis, 45(4), 796–803. 10.1002/jimd.12521

Thompson, J. L. P., Karaa, A., Pham, H., Yeske, P., Krischer, J., Xiao, Y., Long, Y., Kramer, A., Dimmock, D., Holbert, A., Gorski, C., Engelstad, K. M., Buchsbaum, R., Rosales, X. Q., & Hirano, M. (2023). The evolution of the mitochondrial disease diagnostic odyssey. Orphanet J Rare Dis, 18(1), 157. 10.1186/s13023-023-02754-x

Tinker, R. J., Lim, A. Z., Stefanetti, R. J., & McFarland, R. (2021). Current and Emerging Clinical Treatment in Mitochondrial Disease. Mol Diagn Ther, 25(2), 181–206. 10.1007/s40291-020-00510-6

Tsigos, C., Kyrou, I., Kassi, E., & Chrousos, G. P. (2000). Stress: Endocrine Physiology and Pathophysiology. In K. R. Feingold, B. Anawalt, M. R. Blackman, A. Boyce, G. Chrousos, E. Corpas, W. W. de Herder, K. Dhatariya, K. Dungan, J. Hofland, S. Kalra, G. Kaltsas, N. Kapoor, C. Koch, P. Kopp, M. Korbonits, C. S. Kovacs, W. Kuohung, B. Laferrere, M. Levy, E. A. McGee, R. McLachlan, M. New, J. Purnell, R. Sahay, A. S. Shah, F. Singer, M. A. Sperling, C. A. Stratakis, D. L. Trence, & D. P. Wilson (Eds.), Endotext. https://www.ncbi.nlm.nih.gov/pubmed/25905226

Valverde, K. D., McCormick, E. M., & Falk, M. J. (2022). Qualitative exploration of the lived experience of adults diagnosed with primary mitochondrial disease. JIMD Rep, 63(5), 494–507. 10.1002/jmd2.12316

Van den Bergh, O., & Walentynowicz, M. (2016). Accuracy and bias in retrospective symptom reporting. Curr Opin Psychiatry, 29(5), 302–308. 10.1097/YCO.0000000000000267

Zeviani, M., & Carelli, V. (2021). Mitochondrial Retinopathies. Int J Mol Sci, 23(1). 10.3390/ijms23010210

Zilber, S., Yeske, P.E. (2020). Mitochondrial Disease Community Registry: First look at the data, perspectives from patients and families Mitochondrial and Metabolic Medicine, 2. doi:10.9777/mmm.2020.10001

