## Supplemental Figure S2 for "Perceived association of mood and symptom severity in adults with mitochondrial diseases"

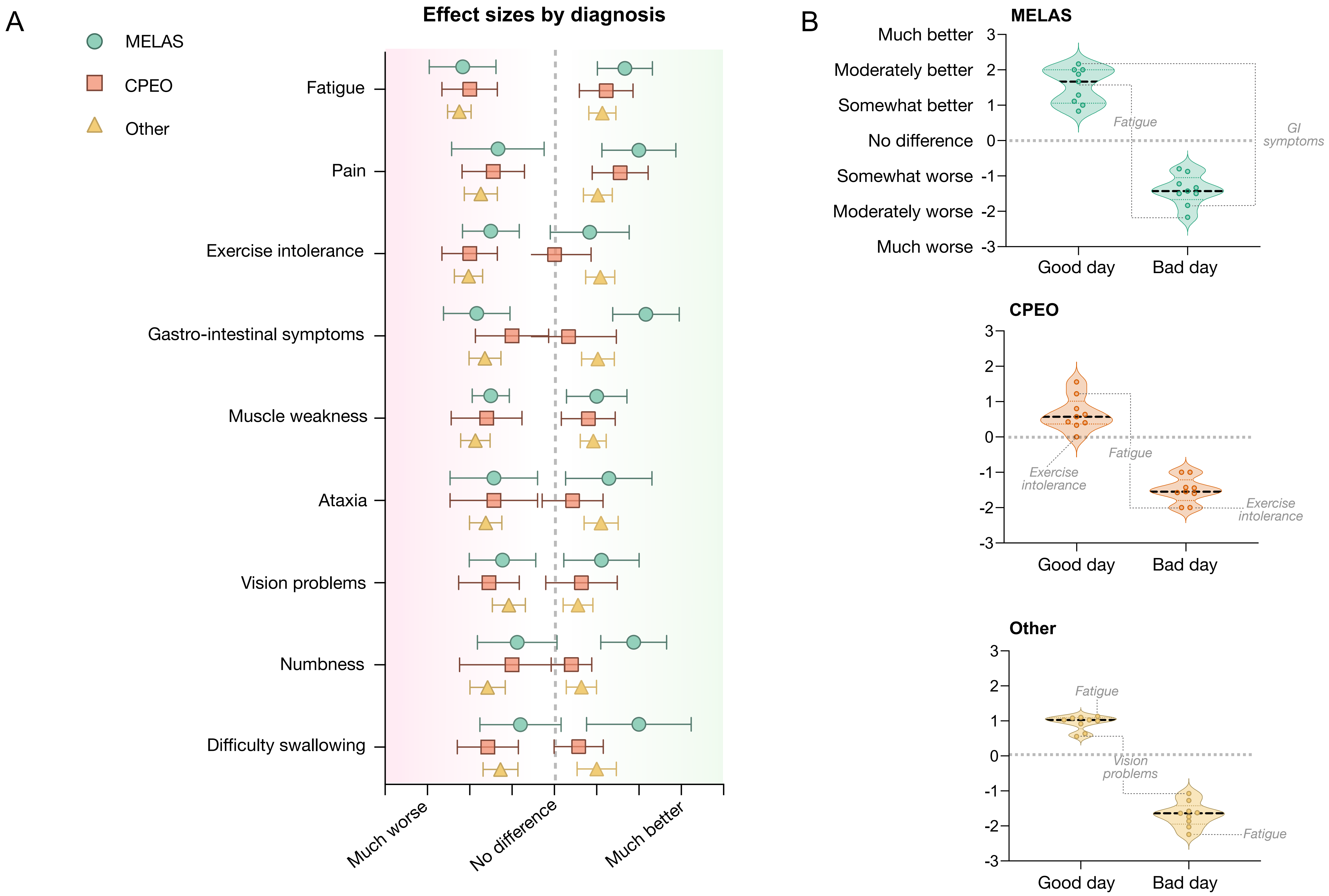


**Supplemental Figure S2. Good/bad day perceived symptom severity scores by clinical diagnoses.** (**A**) Forest plot of a symptoms severity scores (average, 95% CI) on good/bad days by clinical diagnosis (mitochondrial encephalomyopathy, lactic acidosis, and stroke-like episodes (MELAS), chronic progressive external ophthalmoplegia (CPEO), Other). Only symptoms reported by n>3 people were included. (**B**) Difference in perceived symptoms severity computed as a score reflecting the average across all participants (each datapoint represents one symptom), displayed by clinical diagnosis (MELAS, CPEO, Other). Datapoints far away from the dotted line (null hypothesis: no difference) are symptoms perceived to be more strongly to related to mood, in either direction. Bar in violin plots represent the mean of all symptoms.
