## Supplemental Figure S1 for "Perceived association of mood and symptom severity in adults with mitochondrial diseases"

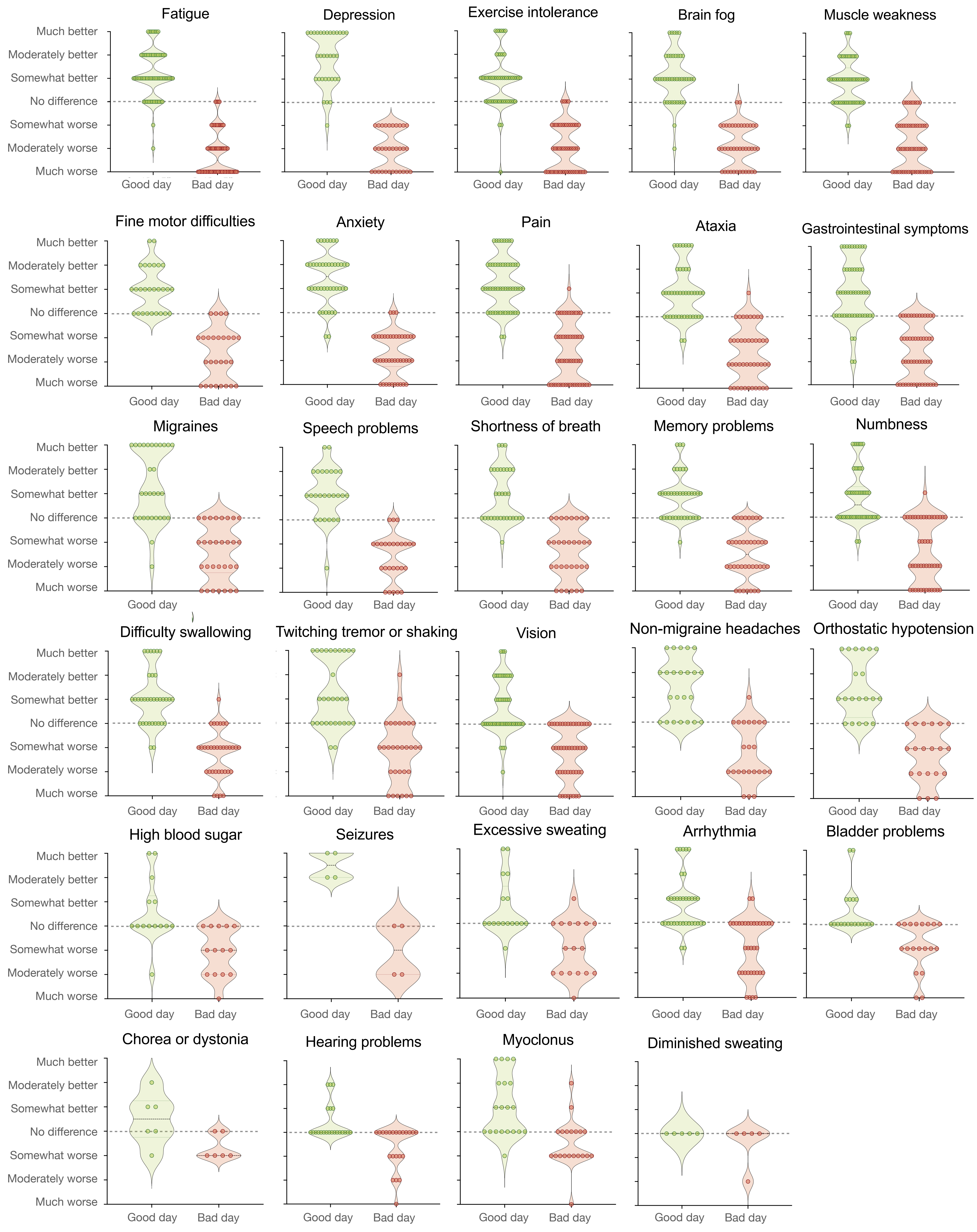


**Supplemental Figure S1. Good/bad day perceived symptom severity.** Perceived symptom severity scores across all participants (each datapoint represents one participant), displayed for each assessed symptom. Datapoints far away from the dotted line (null hypothesis: no difference) are participants who perceived the symptom to be more strongly to related to mood, in either direction. Bar in violin plots represent the mean of all participants.
